# VIBRANT: Vaginal lIve Biotherapeutic RANdomized Trial: A Phase 1 randomized trial of multi-strain vaginal *L. crispatus* live biotherapeutic products in people with bacterial vaginosis

**DOI:** 10.1101/2025.09.18.25336053

**Authors:** Disebo Potloane, Laura Symul, Sinaye Ngcapu, Lara Lewis, Michael France, Laura Vermeren, Joseph Elsherbini, Callin Chetty, Nomfuneko Mafunda, Asthu Mahabeer Polliah, Andile Mtshali, Asavela Kama, Nzuzo Magini, Nireshni Mitchev, Gugulethu Mzobe, Anam Khan, Briah Cooley Demidkina, Miles Goldenberg, Jiawu Xu, Lindsay Rutt, Breanna Shirtliff, Sarah Cook, Meena Murthy, Fatima Hussain, Jo-Ann S. Passmore, Heather B. Jaspan, Brian Kullin, Anna-Ursula Happel, Lenine Liebenberg, Susan Holmes, Douglas S. Kwon, Jacques Ravel, Caroline M. Mitchell

## Abstract

**Introduction:** An optimal vaginal microbiome is typically dominated by beneficial *Lactobacillus* species, whereas bacterial vaginosis (BV) is characterized by high microbial diversity and a paucity of vaginal lactobacilli. High recurrence rates of BV following antibiotic treatment may stem from poor recolonization by protective *Lactobacillus* species post-treatment. This has led to the hypothesis that live biotherapeutic interventions designed to promote *Lactobacillus* dominance could improve BV treatment outcomes.

**Methods:** We conducted a Phase I, double-blind, placebo-controlled randomized trial to evaluate two novel, vaginally delivered live biotherapeutic products (LBP), each containing multiple strains of *Lactobacillus crispatus*. The study was conducted at two sites: one in South Africa, and one in the United States. Eligible participants diagnosed with BV by Nugent score (≥ 7) and Amsel criteria (≥3 out of 4 criteria), first received a course of oral metronidazole and were then randomized equally into one of five arms for 7 days of daily vaginal tablet use: a placebo, a 6-strain LBP (LC106), a 15-strain LBP (LC115), LC106 for 3 days followed by 4 days of placebo, or an unblinded overlap arm in which LC106 was initiated on day 3 of metronidazole treatment. The primary outcomes were safety and detection of any *L. crispatus* strains contained in the products, as determined by metagenomic sequencing within the first weeks of study participation.

**Results:** Across all active arms combined, at least one LBP strain was detected in 66.1% (47/71) of participants at least once in the first five weeks of study participation. Among those with colonization in this period, nearly half (49%, 23/47) remained colonized with LBP strains at 12 weeks, demonstrating durable colonization despite a short initial treatment course. Colonization success was comparable across study arms and sites, although the study was not powered to detect small differences between arms. At both sites, participants were most often colonized by one of three component strains, with no geographic differences in strain colonization observed. Both LBP products were safe, acceptable and well tolerated, with no serious adverse events (AEs) reported. Local/genitourinary AEs occurred most often in the placebo arm.

**Conclusion:** In this Phase 1 study of novel multi-strain *L. crispatus* LBPs, we demonstrated that the products were safe and acceptable, and established durable colonization after a short dosing course in geographically diverse populations. These results provide a foundation for the development of transformational interventions aimed at optimizing the vaginal microbiome.

## Introduction

An optimal vaginal microbiome is characterized by low microbial diversity and dominance by *Lactobacillus* species. This ‘healthy’ state is associated with reduced risk of adverse reproductive health outcomes.^1-4^ In contrast, a non-optimal vaginal microbiome is marked by high diversity and an abundance of obligate and facultative anaerobes, including species in the genera *Gardnerella, Fannyhessea, Prevotella*, and *Mobiluncus*. Women with such microbiota are often diagnosed with bacterial vaginosis (BV), generally have high levels of genital inflammation, and face an increased risk for adverse outcomes such as HIV acquisition and transmission, HPV infection and persistence, cervical dysplasia, miscarriages, and preterm births.^5-10^ Globally, BV affects approximately 30% of women.^11^ It may present with clinical symptoms, including malodorous vaginal discharge, itching, or irritation, or may remain asymptomatic.^12,13^ Since the 1980s, the same two antibiotic classes have remained the standard treatment for clinical BV.^14,15^ While effective in providing short-term symptom relief and reducing BV-associated bacterial burden, recurrences remain unacceptably high. Up to 60% of women experience recurrence within 6 months,^16-18^ likely due to the failure of protective lactobacilli, such as *L. crispatus*, to recolonize the vagina after treatment.^19-21^

Interventions to initiate and promote colonization of the vaginal environment with beneficial *L. crispatus* are therefore a key therapeutic goal. Such strategies may reduce BV recurrence and mitigate the associated adverse reproductive health outcomes. Phase II trials of LACTIN-V, a live biotherapeutic product (LBP) containing a single naturally occurring vaginal strain of *L. crispatus* (CTV-05), showed reduced recurrence rates of clinical BV compared to placebo.^22,23^ However, only 44% of participants in the Lactin-V arm still had detection of the isolate at the final visit, suggesting that inoculation with a single strain of *L. crispatus* may be insufficient for durable colonization of the vagina with beneficial lactobacilli.^24^

To address this limitation, we designed two multi-strain, *L. crispatus*-containing vaginal LBPs using strains isolated from women in the US and South Africa who were stably dominated by *L. crispatus*. The primary objective of this Phase 1 trial was to assess the safety, tolerability and colonization kinetics of these novel, vaginally administered live biotherapeutic products.

## Results

A total of 96 people were randomized, of whom 6 either withdrew after randomization or never used the study product (Figure 1). The modified intent to treat population therefore included 90 participants randomized to five study arms: placebo (N = 19), 7 days of LC106 (LC106-7, N = 21), 3 days of LC106 (LC106-3, N = 15), 7 days of LC106 overlapping with metronidazole (LC106-o, N = 15), and 7 days of LC115 (LC115-7, N = 20). The South African site enrolled the majority of participants (N = 70), evenly distributed across arms (N = 14/arm, Supplemental Figure 1). At the US site, two arms were dropped early due to slow enrollment; one participant was randomized to each of these arms, while the remainder were assigned to placebo (N = 5), LC106 7-day (N = 7), and LC115 (N = 6) (Supplemental Figure 2).

**Figure 1.**
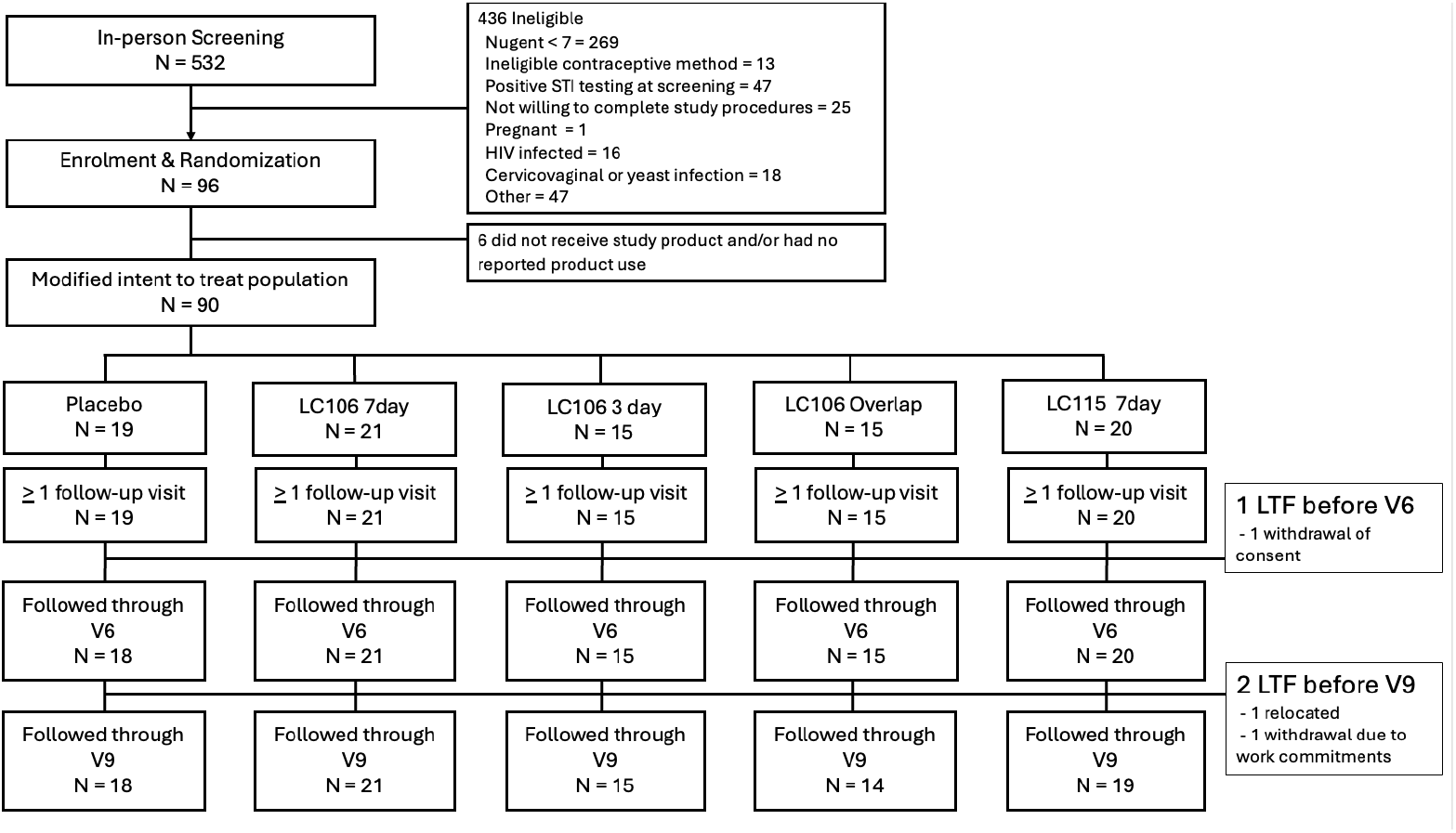
CONSORT diagram representing recruitment, enrollment, randomization and follow up

Demographic characteristics were similar across arms, with notable differences between the two sites in self-identified race/ethnicity and contraceptive type (Table 1, Supplemental Tables 1 and 2). Food insecurity, assessed using the short form of the USDA Food Security Questionnaire,^25^ was more common at the South African site.

**Table 1:** Demographic and behavioral characteristics of participants at enrollment.

|  |  | **Placebo<sup>1</sup>**<br>**N = 19** | **LC106-7<sup>1</sup>**<br>**N = 21** | **LC106-3<sup>1</sup>**<br>**N = 15** | **LC106-o<sup>1</sup>**<br>**N = 15** | **LC115<sup>1</sup>**<br>**N = 20** |
| --- | --- | --- | --- | --- | --- | --- |
| Site | South Africa | 14 (74%) | 14 (67%) | 14 (93%) | 14 (93%) | 14 (70%) |
|  | United States | 5 (26%) | 7 (33%) | 1 (7%) | 1 (7%) | 6 (30%) |
| Age |  | 29 (20-38) | 29 (18-40) | 26 (21-34) | 25 (19-35) | 30(20-40) |
| Race | Asian | 1 (5.3%) | 0 (0%) | 1 (6.7%) | 0 (0%) | 0 (0%) |
|  | Black | 14 (74%) | 18 (86%) | 14 (93%) | 15 (100%) | 18 (90%) |
|  | White | 2 (11%) | 3 (14%) | 0 (0%) | 0 (0%) | 1(5%) |
|  | Other | 1 (5.3%) | 0 (0%) | 0 (0%) | 0 (0%) | 1 (5%) |
|  | Prefer no answer | 1 (5.3%) | 0 (0%) | 0 (0%) | 0 (0%) | 0 (0%) |
| Ethnicity<sup>2</sup> | Not Hispanic | 3 (60%) | 6 (86%) | 1 (100%) | 1 (100%) | 4 (67%) |
|  | Hispanic | 2 (40%) | 1 (14%) | 0 (0%) | 0 (0%) | 2 (33%) |
| Food insecurity | Past 12 months | 7 (37%) | 7 (33%) | 6 (40%) | 5 (33%) | 6 (30%) |
| Contraception | COC | 3 (16%) | 5 (24%) | 1 (6.7%) | 1 (6.7%) | 4 (20%) |
|  | Vaginal ring | 0 (0%) | 1 (4.8%) | 0 (0%) | 0 (0%) | 0 (0%) |
|  | DMPA | 11 (58%) | 10 (48%) | 12 (80%) | 11 (73%) | 11 (55%) |
|  | NET-EN | 2 (11%) | 2 (9.5%) | 2 (13%) | 3 (20%) | 2 (10%) |
|  | Implant | 1 (5.3%) | 0 (0%) | 0 (0%) | 0 (0%) | 0 (0%) |
|  | Lng-IUD | 2 (11%) | 2 (9.5%) | 0 (0%) | 0 (0%) | 2 (10%) |
|  | Other | 0 (0%) | 1 (4.8%) | 0 (0%) | 0 (0%) | 1 (5%) |
| Number of partners past month |  | 1 (1-2) | 1 (0-2) | 1 (0-2) | 1 (0-3) | 1 (0-2) |
| Number of lifetime partners |  | 5 (1-10) | 5 (1-90) | 4 (1-10) | 4 (1-20) | 5 (1-10) |
| Gender partners | No sex | 1 (5%) | 3 (14%) | 1 (6.7%) | 1 (6.7%) | 3 (15%) |
|  | Male only | 18 (95%) | 18 (86%) | 14 (93%) | 14 (93%) | 17 (85%) |
| <sup>1</sup> Median (Min-Max); n (%) |  |  |  |  |  |  |
| <sup>2</sup> Ethnicity percentages only calculated for American participants |  |  |  |  |  |  |
| COC = Combined estrogen/progesterone oral contraceptives; DMPA = Depo medroxyprogesterone acetate; NET-EN = norethisterone enanthate; Implant = etonorgestrel contraceptive implant; Lng-IUD = levonorgestrel containing intrauterine device |  |  |  |  |  |  |

**Table 2:** Proportion of participants at each site who achieved the primary outcome of detection of a live biotherapeutic product strain of *L. crispatus* at > 5% relative abundance at any time during the first 5 weeks. The *p*-values are computed using unconditional exact tests on the data of the two sites combined, comparing rates in each active arm vs placebo and adjusted for multiple comparisons by controlling for the false discovery rate.

|  | Placebo | LC106-7 | LC106-3 | LC106-o | LC115 |
| --- | --- | --- | --- | --- | --- |
| South Africa | N = 14 | N = 14 | N = 14 | N = 14 | N = 14 |
| n (%) with outcome | 0 (0%) | 9 (64%) | 7 (50%) | 7 (50%) | 10 (71%) |
| 95% CI |  | 39%-84% | 27%-73% | 27%-73% | 45%-88% |
| United States | N = 5 | N = 7 | N = 1 | N = 1 | N = 6 |
| n (%) with outcome | 1 (20%) | 6 (86%) | 1 (100%) | 1 (100%) | 6 (100%) |
| 95% CI |  | 49% - 97% | 21% - 100% | 21% - 100% | 61% - 100% |
| RR (95% CI) | Ref | 13.57<br>(1.72 – inf) | 10.13<br>(1.37 – inf) | 10.13<br>(1.37 – inf) | 15.2<br>(2.1 – inf) |
| Adjusted *p* value |  | < 0.001 | < 0.001 | < 0.001 | < 0.001 |

Overall, 66% (47/71; 95% confidence interval [CI]: 54-77%) of women in the active arms achieved the primary outcome: detection of a single LBP strain at ≥ 5% relative abundance, or 2 or more at ≥ 10% total relative abundance as established by metagenomic sequencing in the first five weeks of study participation (Table 2). Detection was highest at week 1 (during dosing) in the LC106-overlap arm, and at week 2 (immediately post-dosing) in the other active arms (Figure 2, Figure 3a). LBP strain detection rates were similar across active arms, though the study was underpowered to detect differences between the different arms (Table 2). At each site, one placebo participant had an LBP strain detected during follow-up, perhaps reflecting misclassification of a native *L. crispatus* strain. Overall, 35% (24/68), 95% confidence interval [CI]: 24-48%) of women in the active arms met the primary outcome at the final 12-week visit.

**Figure 2:**
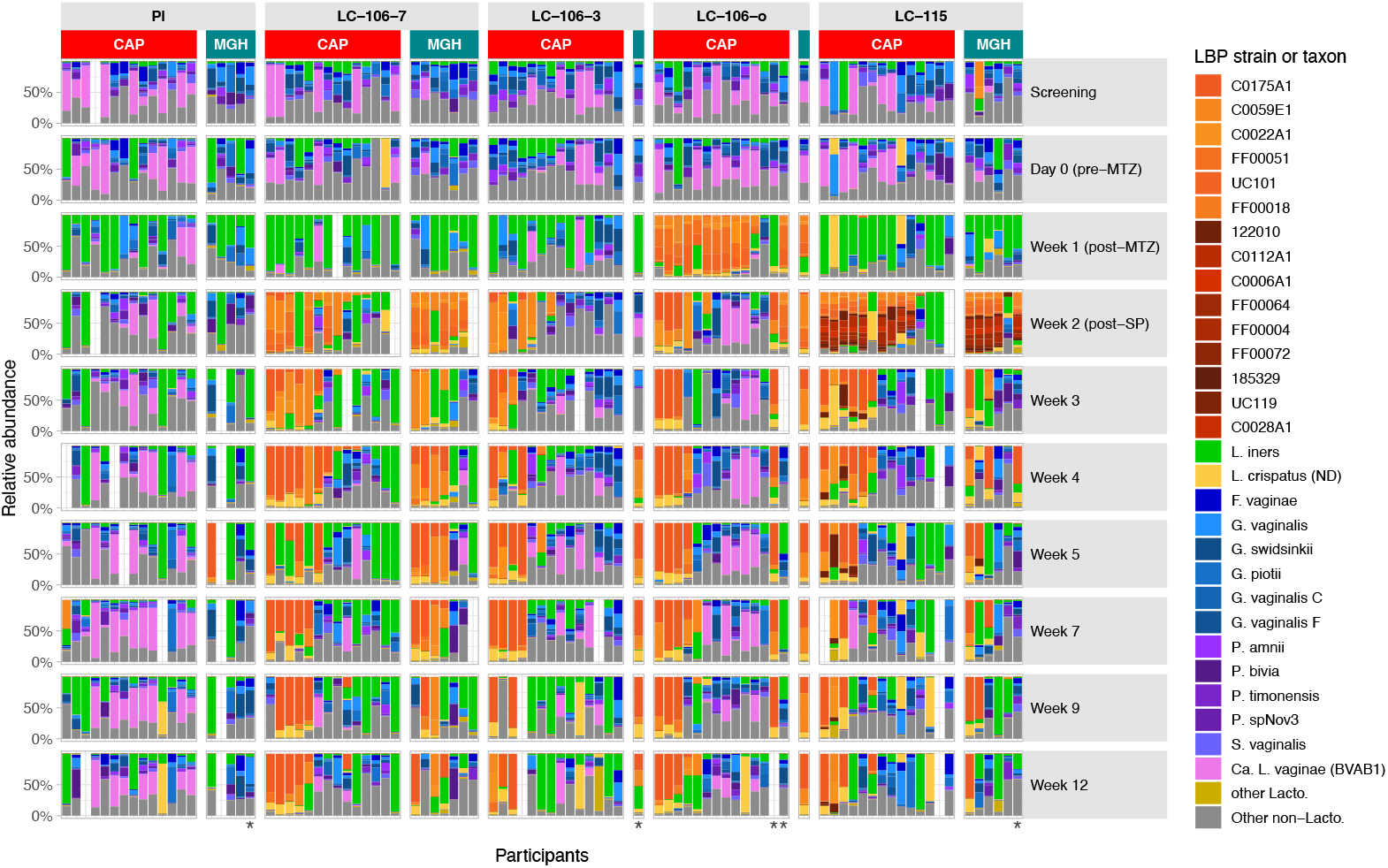
Vaginal bacteria and LBP strains relative abundances estimated by metagenomic sequencing of samples from all clinic visits. Participants are grouped by arm and site (CAP = South African site, MGH = Unites States site), and each participant’s samples are aligned in a single column. Participants marked by an asterisk at the bottom of their column are participants who are part of the modified intent-to-treat population but not the per-protocol population. A white space indicates a missed visit or a sample that failed sequencing. *L. crispatus* strains contained in the live biotherapeutic product are represented in hues of orange, while *L. crispatus* not classified as an LBP strain is yellow.

**Figure 3:**
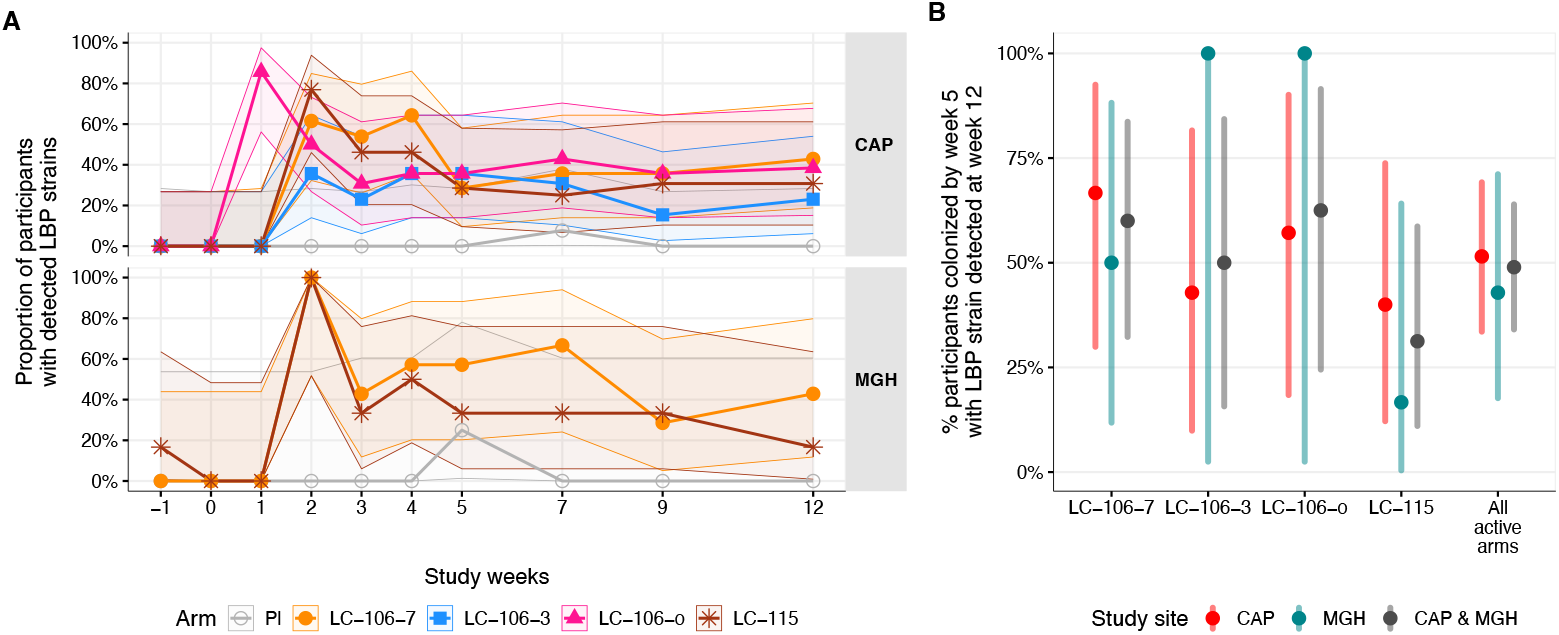
Proportions of (A) all mITT participants with detection of an LBP strain at ≥ 5% relative abundance or a total LBP strain relative abundance ≥ 10% by metagenomic sequencing at each visit, by site (CAP = South African site, MGH = Unites States site; dropped arms at MGH with each a single participant were excluded), and (B) proportion of participants with an LBP strain detected by the metagenomic definition in the first 5 weeks who continue to have an LBP strain detected at week 12.

Among active arm participants who met the primary outcome in the first five weeks, 49% (23/47; 95%CI: 34-64%) remained colonized at week 12, without notable differences between arms (Figure 3b), demonstrating durable colonization despite a short dosing regimen.

At the visit immediately post-dosing in the 7-day arms, some participants had no detectable LBP strains by metagenomics despite reporting using at least 6/7 product doses. Applicator staining for 77 mITT participants who returned their used applicators revealed that 81% (62/77) of participants had high concordance between self-reported use and positive staining. Because 13 mITT participants did not return all applicators, product exposure was further assessed by a post hoc analysis using strain-specific qPCR on daily dosing swabs, applying a strict detection threshold (>10^7^ copies/swab of at least half of the expected LBP strains). With this approach, 90% (64/71) of participants in active arms had detection of LBP strains on at least one dosing day, and 75% (53/71) on at least half of dosing days (Supplemental Figure 3). Among those randomized to seven days of LBP treatment, the mean proportion of days with detection above the threshold was 67% (95% CI: 58%-75%). Participants with detection on at least half of dosing days were more likely to achieve colonization by week 5 than those without (75% [40/53] vs 50% [6/12]).

In the two arms where seven-day dosing began after metronidazole completion, 81% (17/21, LC106-7) and 90% (18/20, LC115-7) of participants had inserted a dose the same day or the day prior to the week 2 visit. Thus, detection at this visit may represent residual study product in the vagina rather than true colonization. A sensitivity analysis using only metagenomic sequencing data from week 3-5 reduced the proportion meeting the primary outcome (LC106-7: 57% vs. 71%; LC115 50% vs. 80%), but colonization rates remained statistically significantly higher than placebo (Supplemental Table 3). Across all visits remote from dosing, when LBP strains were detected by metagenomics, they comprised > 50% of the microbiome in 87% (85/98) of visits in the active arms, suggesting that when retained post dosing, strains typically established dominance.

**Table 3:**
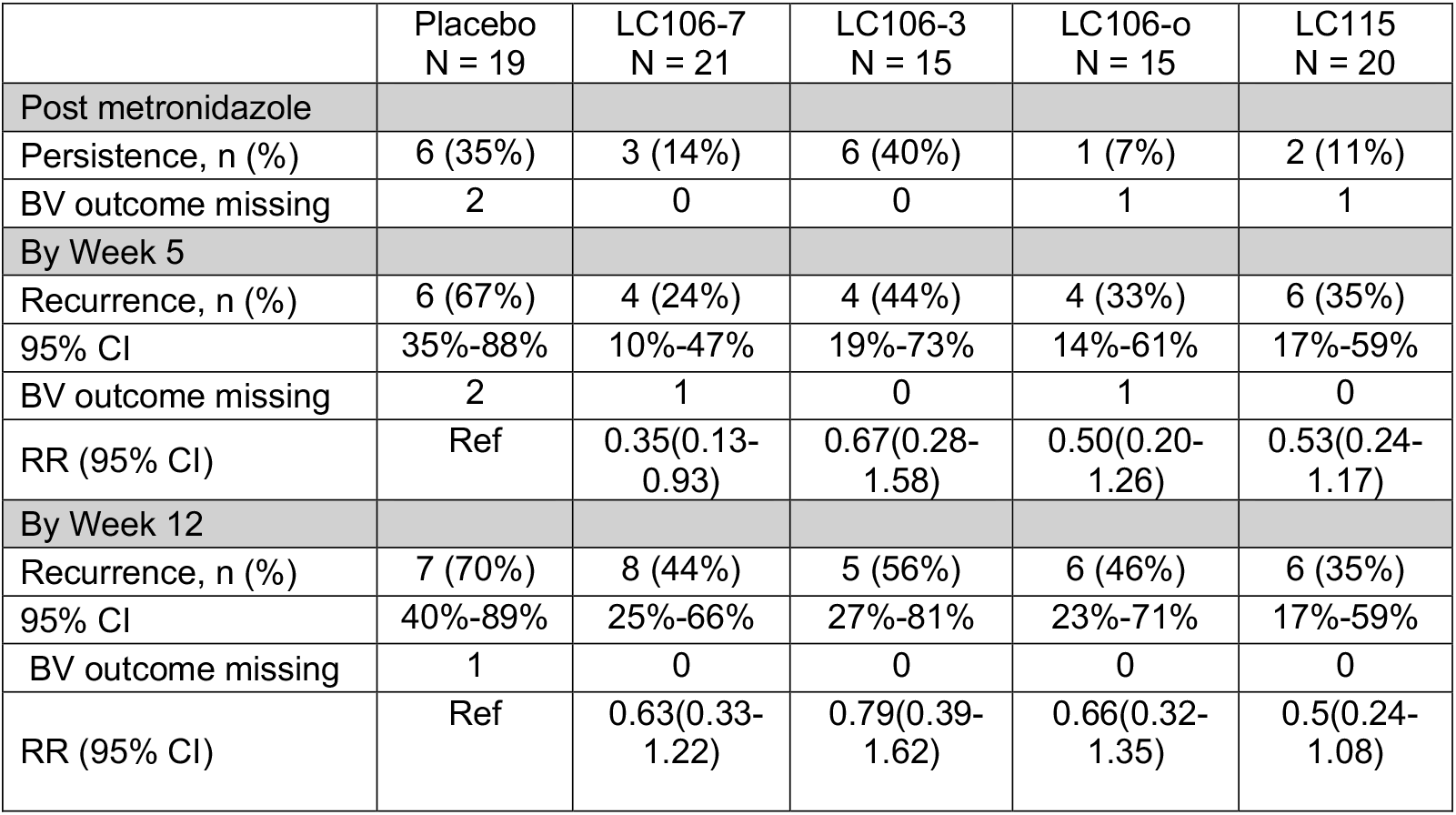
Proportion of people with Nugent score BV (7-10) after metronidazole treatment, by 5 and by 12 weeks.

|  | Placebo<br>N = 19 | LC106-7<br>N = 21 | LC106-3<br>N = 15 | LC106-o<br>N = 15 | LC115<br>N = 20 |
| --- | --- | --- | --- | --- | --- |
| Post metronidazole |  |  |  |  |  |
| Persistence, n (%) | 6 (35%) | 3 (14%) | 6 (40%) | 1 (7%) | 2 (11%) |
| BV outcome missing | 2 | 0 | 0 | 1 | 1 |
| By Week 5 |  |  |  |  |  |
| Recurrence, n (%) | 6 (67%) | 4 (24%) | 4 (44%) | 4 (33%) | 6 (35%) |
| 95% CI | 35%-88% | 10%-47% | 19%-73% | 14%-61% | 17%-59% |
| BV outcome missing | 2 | 1 | 0 | 1 | 0 |
| RR (95% CI) | Ref | 0.35(0.13-0.93) | 0.67(0.28-1.58) | 0.50(0.20-1.26) | 0.53(0.24-1.17) |
| By Week 12 |  |  |  |  |  |
| Recurrence, n (%) | 7 (70%) | 8 (44%) | 5 (56%) | 6 (46%) | 6 (35%) |
| 95% CI | 40%-89% | 25%-66% | 27%-81% | 23%-71% | 17%-59% |
| BV outcome missing | 1 | 0 | 0 | 0 | 0 |
| RR (95% CI) | Ref | 0.63(0.33-1.22) | 0.79(0.39-1.62) | 0.66(0.32-1.35) | 0.5(0.24-1.08) |

Each of the two LBPs contained multiple *L. crispatus* strains from both the US and South Africa (Supplemental Table 4). Most detection events were attributable to three strains: C0022A1, C0059E1, C0175A1 (Figure 4). Detection rates were similar across sites (Figure 5), except for South African strains UC101 and FF00051, which were more frequently detected in US participants immediately post-dosing, though these differences did not persist at later visits and the relative abundance of these strains never exceeded 20% at visits remote from dosing (Figure 4, 5, Supplemental Figure 4).

**Table 4:** Adverse events reported for the safety population during the VIBRANT trial.

|  |  | **Number of events (%of participants with events)** |  |  |  |  |
| --- | --- | --- | --- | --- | --- | --- |
|  |  | Placebo<br>(N=20) | LC106 7<br>(N=21) | LC106 3<br>(N=15) | LC106 overlap<br>(N=15) | LC115 7<br>(N=20) |
| **AEs** | Total | 63(80%) | 53(76%) | 33(73%) | 32(73%) | 46(90%) |
|  | Grade 1 | 33(60%) | 36(57%) | 17(53%) | 13(53%) | 27(65%) |
|  | Grade 2 | 30(65%) | 17(48%) | 16(60%) | 19(60%) | 19(45%) |
|  | Grade 3+ | 0(0%) | 0(0%) | 0(0%) | 0(0%) | 0(0%) |
| **Related AEs** | Total | 11(30%) | 5(19%) | 10(40%) | 4(20%) | 8(25%) |
|  | Grade 1 | 7(25%) | 4(14%) | 8(33%) | 3(20%) | 8(25%) |
|  | Grade 2 | 4(5%) | 1(5%) | 2(13%) | 1(7%) | 0(0%) |
|  | Grade 3+ | 0(0%) | 0(0%) | 0(0%) | 0(0%) | 0(0%) |
| **Local/**<br>**genitourinary AEs** | Total | 39(60%) | 33(52%) | 8(20%) | 9(40%) | 28(55%) |
|  | Grade 1 | 23(35%) | 25(48%) | 6(13%) | 9(40%) | 18(40%) |
|  | Grade 2 | 16(45%) | 8(24%) | 2(13%) | 0(0%) | 10(20%) |
|  | Grade 3+ | 0(0%) | 0(0%) | 0(0%) | 0(0%) | 0(0%) |
| **Severe AEs** | Total | 0(0%) | 0(0%) | 0(0%) | 0(0%) | 0(0%) |

**Figure 4:**
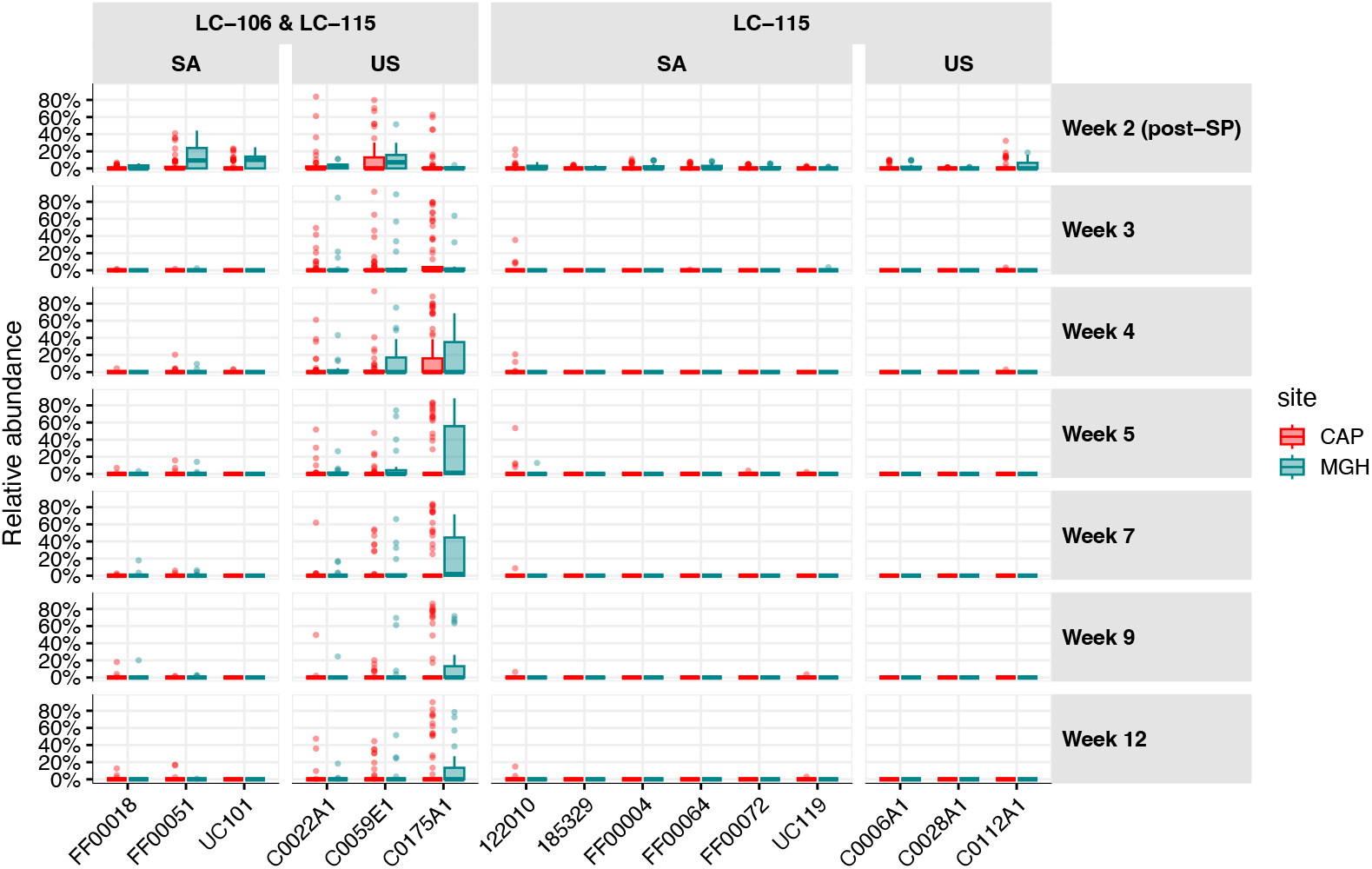
Relative abundance of each strain contained in the live biotherapeutic products at each visit as estimated by metagenomic sequencing. Strains are organized according to geographical origin (SA = South Africa, US = United States), and product(s) in which they are included. Colonization rates are reported separately for each site (CAP = South African site, MGH = Unites States site).

**Figure 5:**
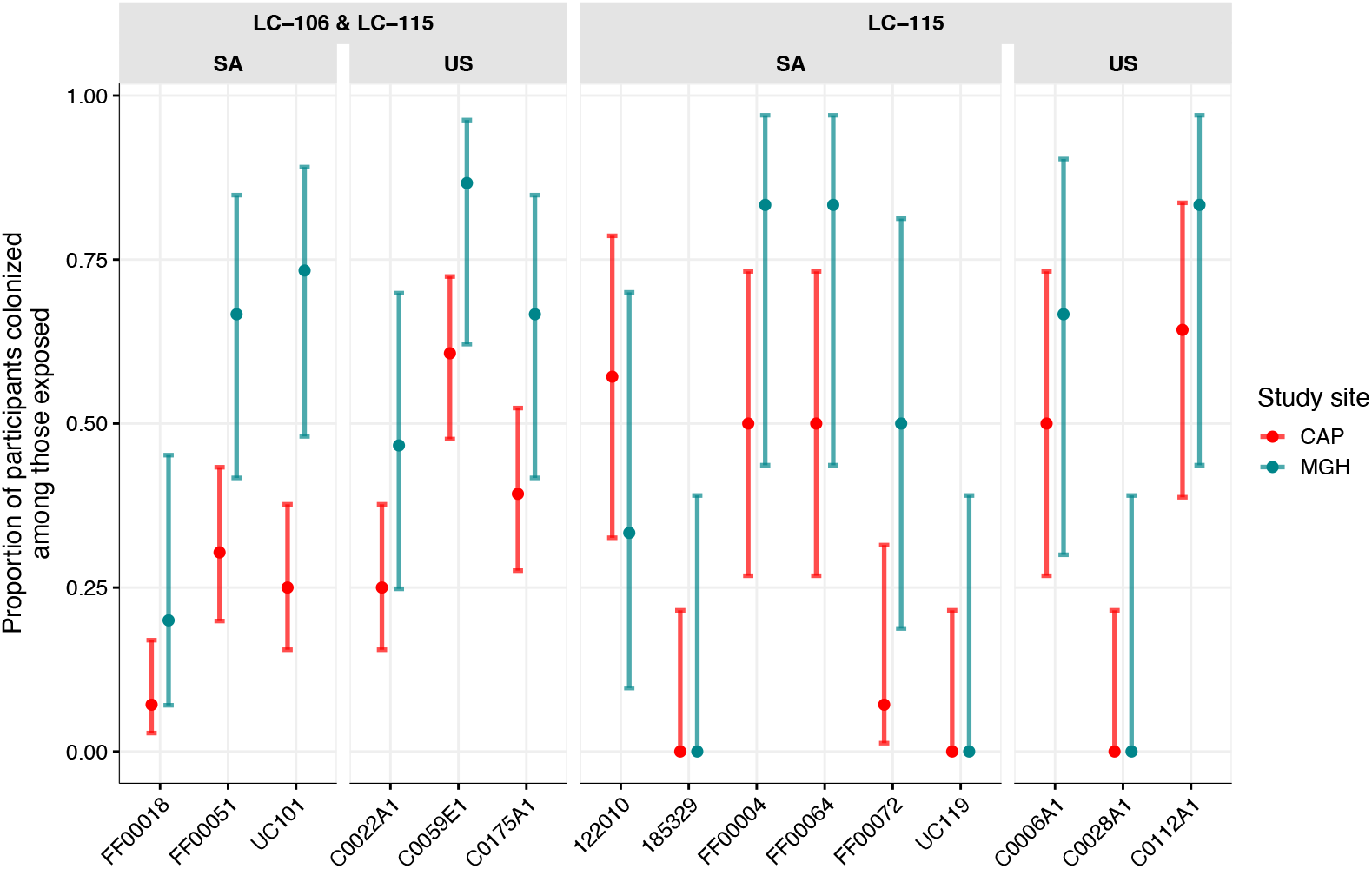
Proportion of women exposed to a specific LBP strain who have that strain detected by metagenomic sequencing at any point during follow up, separated by site. Strains are organized according to geographical origin (SA = South Africa, US = United States), and product(s) in which they are included. Colonization rates are reported separately for each site (CAP = South African site, MGH = Unites States site).

Of the 86 participants with a Nugent score result immediately post-metronidazole treatment, 21%(18/86) had persistent BV (score ≥7). Among these, 12 received an LBP and five (42%) met the primary outcome. The relative risk of BV recurrence among participants receiving an LBP compared to placebo was 0.49 (95% CI: 0.27-0.89) at week 5 and 0.63 (0.38-1.03) at week 12 (Table 3). Among participants with Nugent score < 7 post-metronidazole who received an LBP (n = 57), those achieving the primary outcome by week 5 had a lower probability of BV recurrence over 12 weeks (33%, 13/40; 95%CI 19–49%) than those without colonization (71%, 12/17, 95% CI 44-90%).

Both LBP products were very safe and acceptable. Among the 91 participants included in the safety population, no serious adverse events (AEs) were observed (Table 4). Grade 2 AEs were most frequent in the placebo arm, both overall and for local/genitourinary AEs. Most participants reported willingness to use the product again: 17/19 (89%) placebo, 16/21 (76%) LC106-7, 15/15 (100%) LC106-3, 14/15 (93%) LC106-o, 18/20 (90%) LC115.

## Discussion

We hypothesized that establishment of an *L. crispatus* dominant vaginal microbiome is an essential step in reducing BV recurrence after antibiotic treatment. This randomized, first-in-human, multi-site trial provides the first evidence that a multi-strain exogenous *L. crispatus* LBP can durably engraft—and often persist to 12 weeks—following a short 3–7-day dosing regimen. Over 60% of participants who received LBP demonstrated metagenomic detection of LBP strains, and persistence for up to 12 weeks was observed after as few as three doses.

Importantly, among women cured post-metronidazole, colonization by week 5 was associated with substantially lower BV recurrence over 12 weeks. Findings were consistent across geographically distinct cohorts, suggesting the potential for a single formulation to be effective across populations. Both safety and acceptability were high.

The goal of this LBP is to establish a durable, *L. crispatus*-dominant microbiome, thereby reducing the rates of adverse health outcomes associated with diverse, non*-Lactobacillus-* dominant microbiomes. In this trial, early engraftment was common, and when LBP strains were detected, they typically dominated the community, suggesting that once established, they thrived. Moreover, participants who met our primary outcome of LBP colonization had a lower chance of recurrent BV compared to those who did not, supporting the biological plausibility of a multi-strain *L. crispatus* LBP as an adjunct to antibiotic treatment. However, durability remains the key challenge: by week 12, strains persisted in only 49% of early colonizers, underscoring the need to optimize dose, schedule, and formulation.

The scientific rationale for a multi-strain LBP stems from evidence that in individuals with stable *L. crispatus-*dominated vaginal microbiomes, the metagenome contains more *L. crispatus* genes than can be explained by a single isolate.^26^ We hypothesized that including multiple strains of *L. crispatus* in the LBPs would enhance colonization success by increasing community functional capacity and allowing for host-specific selection, where different women might preferentially support different strains, yielding heterogeneous strain colonization patterns. Contrary to that expectation, three strains consistently emerged as the most frequent colonizers across arms (LC106, LC115) and sites, suggesting broad host compatibility. To extend benefits to a larger population, optimization is still needed – for example, by adding complementary strains or adjusting supporting chemistry/excipients to enhance engraftment and durability.

Our results align with previous studies of the single-strain vaginal *L. crispatus* LBP Lactin-V, which used daily dosing for 5 days followed by twice-weekly maintenance for 4-11 weeks. In those trials, the Lactin-V strain was detectable by qPCR in over half of women one week post-dosing and in less than half 4-12 weeks post-dosing.^22,24^ Similarly, we observed LBP strain detection in over half of participants in the week immediately following treatment using metagenomics, with a reduction to less than half after an additional 7 weeks off product. The risk ratio for BV recurrence in the US Lactin-V Phase 2b study was 0.66 (0.44, 0.87) in the week post-dosing. Notably, our trial demonstrated a relative risk of 0.49 three weeks after dosing, and 0.63 at the three-month follow-up visit. These rates were achieved without maintenance dosing, suggesting that durable colonization may occur early or not at all, and that a short dosing course may be sufficient.

Several limitations warrant consideration. Due to the modest sample size, the study was underpowered to detect small differences between active study arms, limiting conclusions about the optimal dosing strategy or LBP composition. Adherence was measured by participant report and examination of returned tablet packages and applicators, which may not fully capture actual use, However, post-hoc analysis of strain-specific qPCR on daily swabs collected during dosing showed good agreement between self-reported adherence and observed exposure to the LBP strains. Site differences also influenced study conduct: in the US, telephone pre-screening was used, whereas in South Africa, screening was conducted exclusively in person. Stringent inclusion criteria also affected enrollment differently at the two sites: in the US, there was a higher-than-expected preference for non-hormonal contraceptive methods, while in South Africa, a high rate of Amsel-negative, but Nugent-positive BV slowed accrual, necessitating study protocol amendments.

## Conclusion

In this Phase 1 study of novel multi-strain *L. crispatus* LBPs, we demonstrated that these products were safe, acceptable, and capable of establishing durable colonization after a short dosing regimen in geographically diverse populations. These findings lay the foundation for the development of transformational interventions to optimize the vaginal microbiome.

## Methods

### Study design and participants

From February 9, 2024 through February 19, 2025 we conducted a two-site randomized, controlled Phase 1 trial of two multi-strain *L. crispatus* vaginal live biotherapeutic products in Vulindlela, KwaZulu Natal, South Africa and Boston, Massachusetts, United States. The protocol was reviewed and approved by SAHPRA (20230615) and FDA (IND 029629), as well as ethics committees at both sites (BREC/00005620/2023; MGB IRB 2023P001035). The study was registered on clinicaltrials.gov (NCT06135974).

Participants were recruited through in-person community outreach teams (South Africa), online and physical advertisements (US), direct recruitment through the electronic health record (US), and local clinics (both sites). At the US site, telephone pre-screening was conducted to reduce the number of in-person screening visits. At screening visits, cis-gender women 18-40 underwent eligibility testing. Inclusion criteria included a diagnosis of bacterial vaginosis (BV), and negative testing for yeast (by microscopy), *Neisseria gonorrhoeae, Chlamydia trachomatis, Trichomonas vaginalis* (by NAAT), as well as negative serology for syphilis and HIV. Exclusion criteria included pregnancy, significant medical history, use of antibiotics or probiotics in the past 30 days. Complete inclusion and exclusion criteria are detailed in the study protocol.^27^

Initially, BV was diagnosed at both sites using both Amsel criteria (≥ 3/4)^28^ and Nugent score 7-10^29^ at both sites. However, at the South African site many participants met Nugent criteria but not Amsel criteria. A review of the literature indicated that this discrepancy is common in microbiome studies in African populations,^30^ and so the Amsel criteria were dropped at the South African site in July 2024.

Initially, all participants were required to use either continuous combined oral contraceptives or injectable progestin-only contraceptives to suppress menses. However reluctance to use hormonal contraception or current use of an IUD were two of the primary reasons for exclusion during pre-screening in the US. In August 2024 the protocol in the US was amended to allow participants to use any form of contraception.

### Interventions

All participants received oral metronidazole, taken twice daily for 7 days. The dosage differed between the sites according to local standards of care: 500mg twice daily in US, and 400mg twice daily in South Africa. Participants were randomized to one of five arms: 7 days of vaginal placebo tablet following metronidazole, 7 days of LC106, a six-strain *L. crispatus* live biotherapeutic vaginal tablet, following metronidazole (LC106-7), 3 days of LC106 + 4 days of placebo following metronidazole (LC106-3), 7 days of LC106 starting on day 3 of metronidazole (overlap arm, LC106-o), and 7 days of LC115, a fifteen-strain *L. crispatus* live biotherapeutic vaginal tablet following metronidazole (LC115-7). Due to slow accrual at the US site, LC106-3 and the LC106 overlap arms were dropped in May 2024, and planned enrollment for the South African site was increased.

### Randomization and blinding

An unblinded statistician created randomization lists stratified by site. Randomization was performed at the time of enrollment (*i*.*e*., once eligibility was confirmed) using sequentially numbered, opaque envelopes. The “overlap” arm was unblinded, but all other arms received blinded study product, dispensed in identical-appearing boxes. When two arms were dropped at the US site, the unblinded statistician informed the study team which randomization envelopes to remove.

### Procedures

Participants meeting eligibility criteria after screening returned for an enrollment visit, were dispensed oral metronidazole and were randomized. Study product was dispensed at week 1 visit (after metronidazole completion) or at enrollment for those in the overlap arm.

Participants in all but the overlap arm initiated study product within 24 hours of completing metronidazole. Follow-up included weekly in-person visits until week 5, and additional visits at week 7, 9 and 12. At each in-person visit participants self-collected vaginal swabs. Blood was drawn for testing for syphilis and HIV serology testing at screening, week 5 and week 12.

Additionally, vaginal swabs were collected for *N. gonorrhoeae, C. trachomatis and T. vaginalis* testing at the same visits. Between enrollment and the week 5 visits, participants completed daily diaries documenting product use, bleeding and sexual activity and self-collected a single vaginal swab which was placed in a nucleic acid stabilizing buffer stored at room temperature and returned to the study team at weekly visits.

### Outcomes

The primary outcomes for the study were 1) vaginal detection of any single strains included in the LBP at ≥ 5% relative abundance, or any combination of LBP strains at ≥ 10% relative abundance using metagenomic sequencing at any point prior to the 5-week study visit and 2) safety, as measured by reported adverse events. If participants missed a visit or if metagenomic sequencing could not be done for a sample, we assumed absence of colonization at that time point (worst-case assumption). Secondary outcomes included detection of the LBP strains between week 5 and week 12, recurrence of BV by Amsel criteria and/or Nugent score, the proportion of people with *L. crispatus*-dominance (> 50% relative abundance), and acceptability of the study product.

### Sample processing

Vaginal flocked swabs were placed in cryovials with either 1 mL C2 buffer, 1 mL 10% glycerol/thioglycolate solution, or no liquid and were stored at -80C until use.

Polyester swabs were collected for Amsel and Nugent assessment. Nugent scoring was performed by at least two independent readers, and if scores differed by more than 2 points or spanned diagnosis categories a third reader was used to adjudicate the score. Each site cross-read slides from the other site to ensure that scoring consistency.

### Measurement of adherence to metronidazole and LBP

Adherence to both metronidazole and study product was assessed using: 1) participant daily diary, 2) self-report at the weekly in-person visits, 3) vaginal applicator staining with 0.05% methylene blue solution for study product.^31,32^

### DNA extraction

Vaginal swab specimens collected into C2 buffer were transferred to 96-well plates for automated DNA extraction using the MagAttract PowerMicrobiome DNA/RNA EP Kit (Qiagen, Germantown, MD, USA) on a custom protocol automated on a Microlab STAR robotic platform (Hamilton, Reno, NV, USA). Mechanical lysis was performed on a TissueLyser II (Qiagen) at 20 Hz for 20 minutes to lyse both gram-positive and gram-negative bacteria. Each extraction run included negative controls (nuclease-free water and unused swabs in C2 buffer) and positive controls consisting of two defined mock communities: Mock1 (all 15 *Lactobacillus crispatus* strains plus a standardized vaginal microbial community) and Mock2 (the 15 *L. crispatus* and selected bacterial isolates). DNA was purified via magnetic bead separation, eluted in nuclease-free water and quantified using a Qubit fluorometer (Thermo Fisher Scientific, USA) to confirm sufficient yield and quality for downstream 16S rRNA gene sequencing, qPCR and metagenomic sequencing.

### Metagenomic sequencing & bioinformatic processing

The shotgun metagenomic sequence libraries were constructed from the extracted DNA using Illumina Nextera XT kits (Illumina Inc., California USA), according to manufacturer’s instructions. The resulting libraries were sequenced on an Illumina NovaSeq 6000 (150 bp paired end mode, targeting 45 million read pairs per sample) at Maryland Genomics (https://marylandgenomics.org/). Human reads were detected and removed from all metagenomic sequencing data using BMTagger and the GRCh38 reference genome^33^, and remaining reads were quality filtered using fastp^34^ (v0.22.0, sliding window: size 4 bp, Q12, minimum read length:55 bp). The reads were then mapped to the VIRGO2 non-redundant gene catalog to establish taxonomic composition^35^ (v1, default settings). The relative abundances of each taxon were computed as the proportion of the gene-length-adjusted number of reads assigned to that taxon over the sum of all gene-length-adjusted reads assigned to bacterial content (*i*.*e*., excluding reads assigned to the human host genome or to the genomes of non-bacterial microbiota organisms). We next used kSanity to detect and quantify the abundance of the *L. crispatus* strains that comprise LC106 and LC115^36^ (v1, k=55). The output of kSanity reports the abundance of each LBP strain, relative to the abundance of all *L. crispatus*. The LBP strain abundances were then multiplied by the overall relative abundance of *L. crispatus*, as estimated by VIRGO2, to establish their relative abundance in the overall community.

### Strain-specific qPCR assays

Detection and quantification of LBP strains was also performed using strain-specific qPCR assays. A total of 15 assays were developed, each targeting a unique gene region of an individual *L. crispatus* strain present in the LBPs (Supplementary Table 5). The 15 assays were multiplexed in groups of three using three different fluorophores (Cy5, HEX or FAM). The qPCR reactions included the following components: 0.5µL of each primer mix (18µM forward primer, 18µM reverse primer, 5µM probe), 5µL of PrimeTime qPCR master mix (IDT, New Jersey, USA), 2µL of DNA template, and 1.5µL of water (10µL total reaction volume). Reactions were performed on a CFX-384 Touch Real-Time PCR Detection System (Bio-Rad Laboratories, Pennsylvania USA), in triplicate with a gDNA standard curve spanning from 10^1^ to 10^7^ copies per µL. Thermocycling included an initial 10min of denaturing at 95°C followed by 40 cycles of 95°C for 15s and 62°C for 1 minute.

### Statistical analysis

#### Sample size calculation

We assumed that at most 10% of participants in the placebo arm would have detectable LBP strains. We defined a clinically significant difference between arms as a ≥ 50% absolute difference in LBP strain detection between any active arm and placebo. With 10 participants per arm, the trial had 82.3% power to detect a difference of 60% in the colonization proportion between an active arm and placebo using a one-sided Fisher’s Exact test with a significance threshold of 0.05 (PASS 2020). Our initial goal was to recruit sufficient numbers at each site to allow within-site analyses, however due to slow accrual, the US site did not achieve 10 people per arm.

#### Analysis populations

For the assessment of product safety we included data from all randomized participants who met inclusion/ exclusion criteria and received at least one LBP dose and who had any post-LBP data available. Participants were analyzed according to treatment received. For analyses of colonization we used a modified intent-to-treat (mITT) population that included all randomized participants who met inclusion/exclusion criteria, used at least one dose of study product, and returned for at least one follow-up visit. The per protocol (PP) analyses included all randomized people who met inclusion/ exclusion criteria, did not meet replacement criteria, completed at least 80% of study product doses (by self-report and/or applicator staining), and had at least one follow-up visit prior to week 7. Participants were analyzed in the groups to which they were assigned. Four participants met replacement criteria, of whom two were replaced. All four were included in the mITT population, but only the final two were included in the PP population.

#### Primary outcome testing

One-sided unconditional exact test (Fisher-adjusted method)^37^ was used to test whether the colonization proportion in each of the active arms was greater than in the placebo arm (data from both sites were considered simultaneously). Multiple testing was accounted for by adjusting the *p*-value to control for the false-discovery rate using the Benjamini-Hochberg procedure.^38^ Confidence intervals for proportions (primary outcomes and all other proportions reported here) were computed using the “score” method.^39^

#### Secondary outcome testing

For the analysis of BV recurrence, risk ratios and 95% Wald confidence intervals were calculated. Since adjustment for multiple testing for secondary objectives was not defined a priori, adjusted for multiplicity was not performed and confidence intervals should not be used to infer definitive effects.

#### Post-hoc analyses

Our daily qPCR data was generated after unblinding, however the technicians and bioinformaticians processing the samples and data were blinded to participant/sample assignment. For the mean proportion of days exposed, confidence intervals were calculated using Student’s method.

#### Statistical test used in Table 1

Continuous variables summarized by median and range were compared across study arms using the Kruskal-Wallis rank sum test. Categorical variables were analyzed using either Fisher’s exact test (for small cell counts) or Pearson’s Chi-squared test (when expected frequencies were sufficient), depending on the distribution of the data.

## Data Availability

All data produced in the present study are available upon reasonable request to the authors

## Funding

This project was funded by the Gates Foundation (INV-019055 and INV-037901 to CM, and INV-037902 To DP)

## Acknowledgments

We thank Lara Wautier (UCLouvain) and Precious Radebe (CAPRISA) for their help with the management and formatting of clinical data.

All sequencing data were generated by Maryland Genomics, Institute for Genome Sciences, University of Maryland School of Medicine, Baltimore, MD. We want to thank Mike Humphrys and Lisa Bilski for their contribution to preparing and dispatching sampling kits to the clinical sites.

## Author contributions

Conceptualization: DP, JAP, DSK, JR, CMM

Developed the study protocol: DP, LS, SN, LL, CC, NoM, AMP, JAP, AUH, SH, DSK, JR, CMM

Provided isolates for the LBP: SN, JAP, BK, HJ, AUH, DSK, JR

Conducted the clinical trial: DP, CC, NoM, AMP, AK, BCD, CMM

Processed and evaluated trial samples: SN, AM, AK, NzM, NiM, GM, MG

Generated laboratory data for trial samples: SN, AMP, AM, AK, NzM, NiM, GM, AK, BCD, MG, JX, LR, BS, SC, LL

Performed analysis: LS, LL, MF, LV, JE, AK

Writing of initial draft: DP, LS, LL, LV, CMM

Review of final draft: All authors

## Competing Interests

CMM has been a consultant for Freya Biosciences and serves on the scientific advisory board of Ancilia Biosciences and Concerto Biosciences. CM has a financial interest in Ancilia Biosciences, a company developing a new class of Live Biotherapeutics and other bacterial products. Dr. Mitchell’s interests were reviewed and are managed by MGH and Mass General Brigham in accordance with their conflict-of-interest policies. J.R. is co-founder of LUCA Biologics, a biotechnology company focusing on translating microbiome research into live biotherapeutic drugs for women’s health

## Notes

### Clinical Trial

NCT06135974

### Funding Statement

This project was funded by the Gates Foundation (INV-019055 and INV-037901 to CMM, and INV-037902 To DP)

### Author Declarations

The study was reviewed and approved by institutional review boards at the University of KwaZulu Natal (BREC/00005620/2023) and Mass General Brigham (MGB IRB 2023P001035)

### Summary of Updates

We added two authors who were inadvertantly left off. We added a specific statement about the rate of detection of LBP strains at 12 weeks to the results section.

