## Supplemental Material for "VIBRANT: Vaginal lIve Biotherapeutic RANdomized Trial: A Phase 1 randomized trial of multi-strain vaginal *L. crispatus* live biotherapeutic products in people with bacterial vaginosis"

**Supplemental Figure 1**: CONSORT for the South African site alone

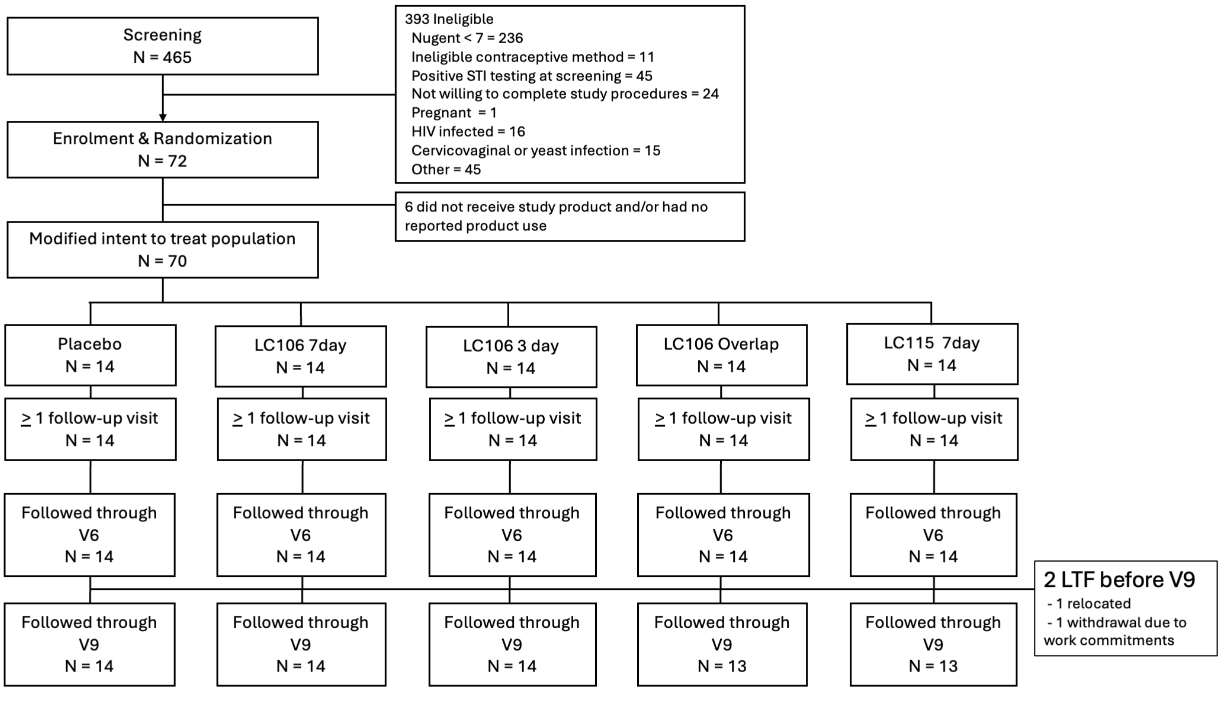

**Supplemental Figure 2**: CONSORT for the US site alone

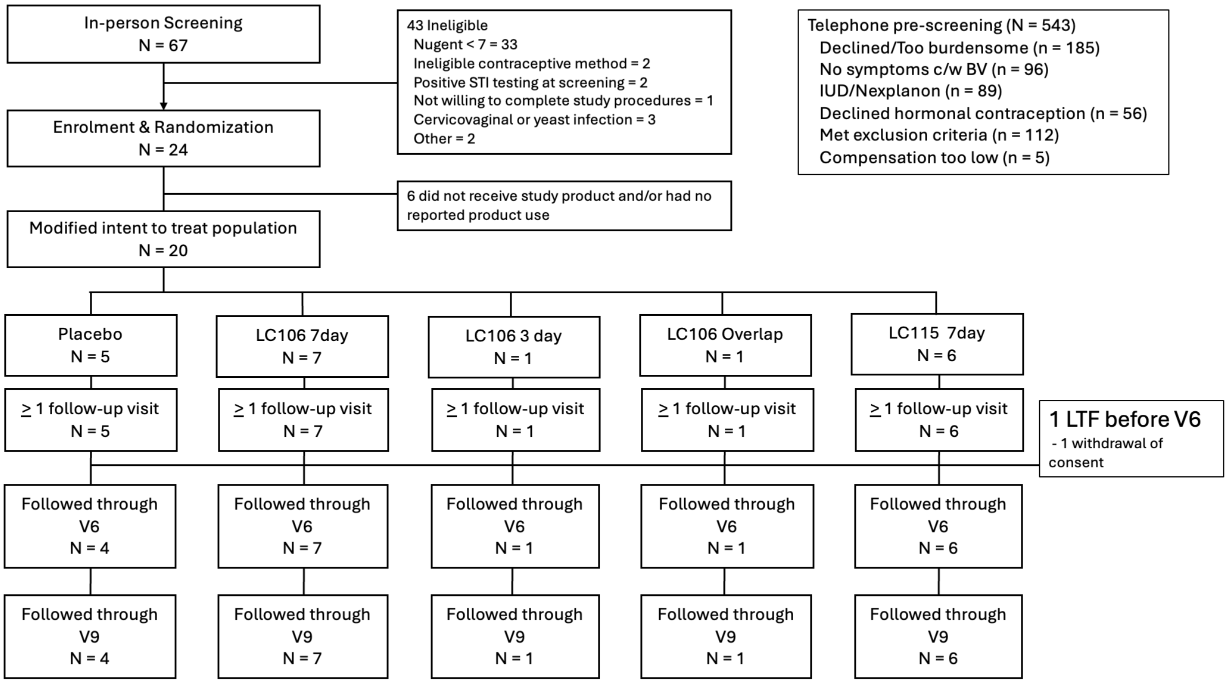

**Supplemental Figure 3:** Proportion of product used by participants in each active arm, according to the daily qPCR definition (detecting at least half of the expected LBP strains at > $10^7$ copies/swab)

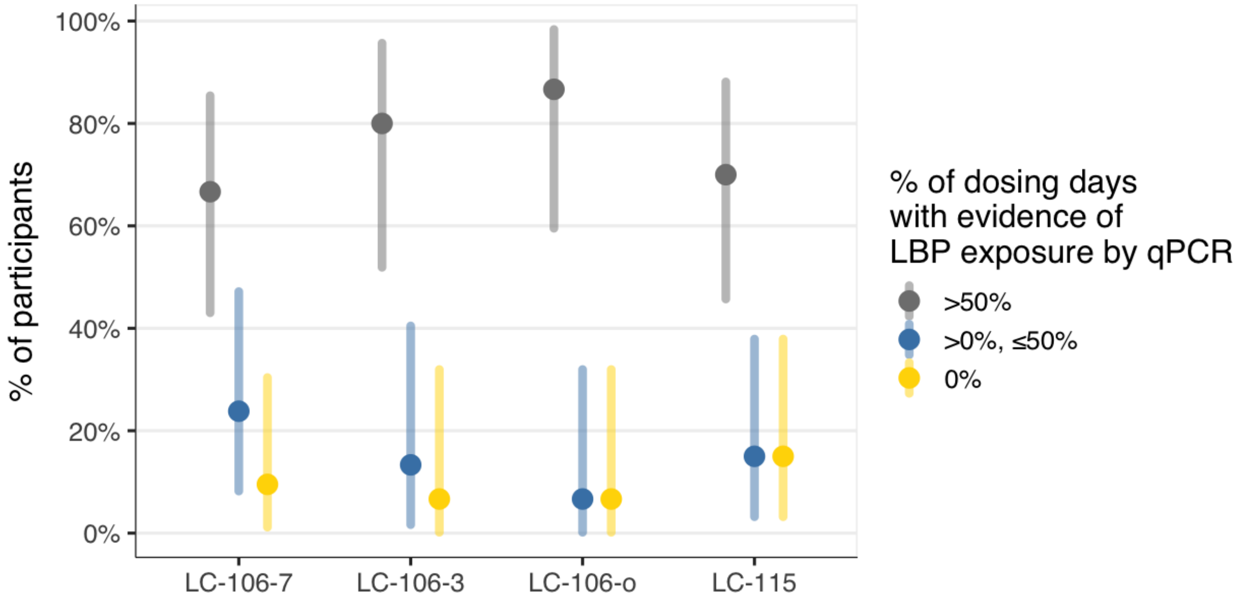

**Supplemental Figure 4:** Proportion of women exposed to a specific strain who have that strain detected by metagenomic sequencing at visit 3-5, separated by site. Strains are organized according to geographical origin (SA = South Africa, US = United States), and product(s) in which they are included. Colonization rates are reported separately for each site (CAP = South African site, MGH = Unites States site).

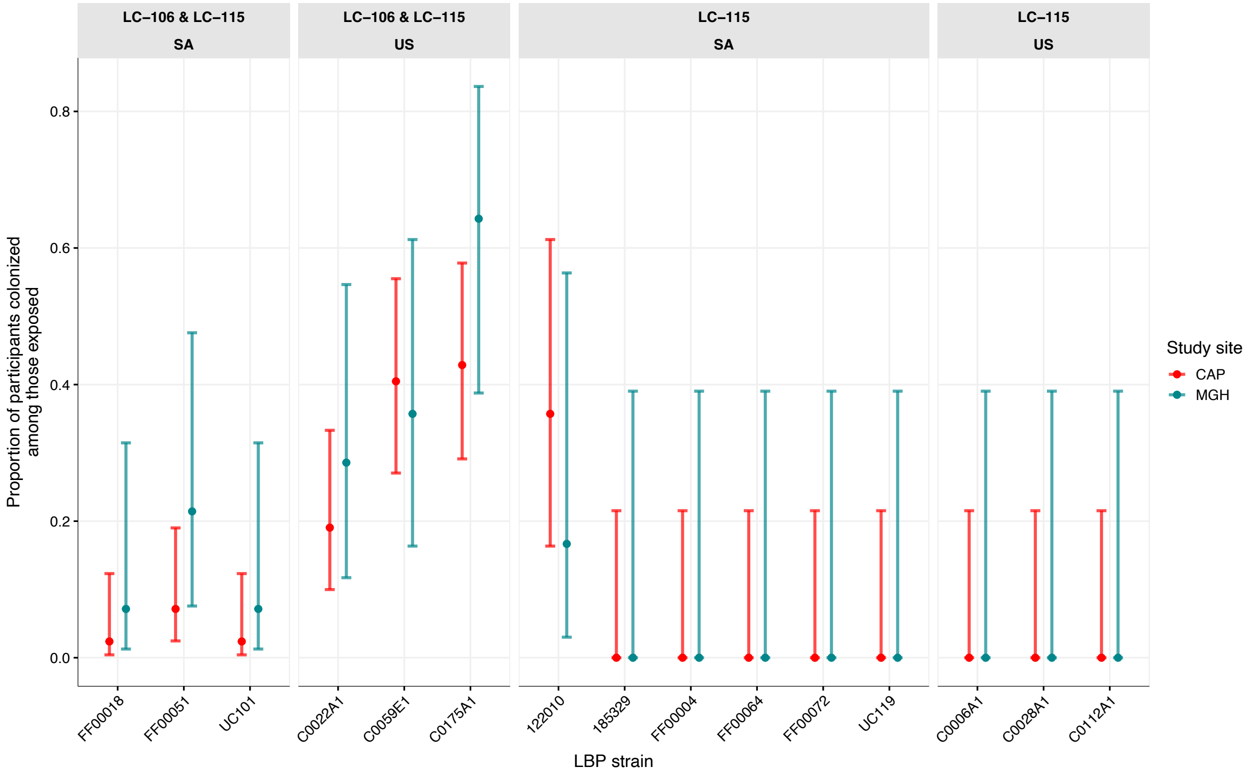

**Supplemental Table 1:** Demographic and behavioral characteristics of participants at enrollment, by arm and by site

| | | Placebo[1] | LC106-7[1] | LC106-3[1] | LC106-o[1] | LC115[1] |
|---|---|---|---|---|---|---|
| South Africa | | N = 14 | N = 14 | N = 14 | N = 14 | N = 14 |
| Age | | 28 (20-36) | 26 (18-36) | 26 (21-34) | 26 (19-35) | 27 (20-37) |
| Race | Black | 14 (100%) | 14 (100%) | 14 (100%) | 14 (100%) | 14 (100%) |
| Food insecurity | Past 12 months | 7 (37%) | 5 (36%) | 6 (43%) | 5 (36%) | 5 (36%) |
| Education | Primary | | | | | |
| | Secondary | | | | | |
| | Tertiary | | | | | |
| Contraception | COC | 1 (7.1%) | 2 (14%) | 0 (0%) | 0 (0%) | 2 (14%) |
| | DMPA | 11 (79%) | 10 (71%) | 12 (86%) | 11 (79%) | 10 (71%) |
| | NET-EN | 2 (14%) | 2 (14%) | 2 (14%) | 3 (21%) | 2 (14%) |
| | Implant | 1 (5.3%) | 0 (0%) | 0 (0%) | 0 (0%) | 0 (0%) |
| | Lng-IUD | 2 (11%) | 2 (9.5%) | 0 (0%) | 0 (0%) | 2 (10%) |
| | Other | 0 (0%) | 1 (4.8%) | 0 (0%) | 0 (0%) | 1 (5%) |
| Number of partners past month | | 1 (1-2) | 1 (1-2) | 1 (0-2) | 1 (0-3) | 1 (1-2) |
| Number of lifetime partners | | 5 (1-10) | 4 (1-7) | 4 (1-10) | 6 (1-20) | 4 (1-9) |
| Gender partners | No sex | 1 (7.1%) | 2 (14%) | 1 (6.1%) | 1 (7.1%) | 2 (14%) |
| | Male only | 13 (93%) | 12 (86%) | 13 (93%) | 13 (93%) | 12 (86%) |
| United States | | N = 5 | N = 7 | N = 1 | N = 1 | N = 6 |
| Age | | 32 (26-38) | 33 (24-40) | 25 (25-25) | 35 (35-35) | 33 (22-40) |
| Race | Asian | 1 (20%) | 0 (0%) | 1 (100%) | 0 (0%) | 0 (0%) |
| | Black | 0 (0%) | 4 (57%) | 0 (0%) | 1 (100%) | 4 (67%) |
| | White | 2 (40%) | 3 (43%) | 0 (0%) | 0 (0%) | 0 (0%) |
| | Other | 1 (20%) | 0 (0%) | 0 (0%) | 0 (0%) | 1 (17%) |
| | Prefer no answer | 1 (20%) | 0 (0%) | 0 (0%) | 0 (0%) | 0 (0%) |
| Ethnicity* | Not Hispanic | 3 (60%) | 6 (86%) | 1 (100%) | 1 (100%) | 4 (67%) |
| | Hispanic | 2 (40%) | 1 (14%) | 0 (0%) | 0 (0%) | 2 (33%) |

| | | | | | | |
|---|---|---|---|---|---|---|
| Food insecurity | Past 12 months | 0 (0%) | 2 (29%) | 0 (0%) | 0 (0%) | 1 (17%) |
| Contraception | COC | 2 (40%) | 3 (43%) | 1 (100%) | 1 (100%) | 2 (40%) |
| | Vaginal ring | 0 (0%) | 1 (14%) | 0 (0%) | 0 (0%) | 0 (0%) |
| | DMPA | 0 (0%) | 0 (0%) | 0 (0%) | 0 (0%) | 1 (17%) |
| | Implant | 1 (20%) | 0 (0%) | 0 (0%) | 0 (0%) | 0 (0%) |
| | Lng-IUD | 2 (40%) | 2 (29%) | 0 (0%) | 0 (0%) | 2 (33%) |
| | Other | 0 (0%) | 1 (14%) | 0 (0%) | 0 (0%) | 1 (17%) |
| Number of partners past month | | 1 (1-2) | 1 (0-2) | 1 (1-1) | 1 (1-1) | 1 (0-2) |
| Number of lifetime partners | | 9 (7-10) | 25 (6-90) | 7 (7-7) | 10 (10-10) | 7 (4-10) |
| Gender partners | No sex | 0 (0%) | 1 (14%) | 0 (0%) | 0 (0%) | 1 (17%) |
| | Male only | 5 (100%) | 6 (86%) | 1 (100%) | 1 (100%) | 5 (83%) |

[1]Mean (Min-Max); n (%)
COC = Combined estrogen/progesterone oral contraceptives; DMPA = Depo medroxyprogesterone acetate; NET-EN = norethisterone enanthate; Implant = etonorgestrel contraceptive implant; Lng-IUD = levonorgestrel containing intrauterine device

**Supplemental Table 2:** Variables that are significantly different between South African (CAPRISA) and US (MGH) participants

| | | CAPRISA (N = 70) | MGH (N = 20) | P value |
|---|---|---|---|---|
| Race | Asian | 0 | 2 (10%) | < 0.001 |
| | Black | 70 (100%) | 9 (45%) | |
| | White | 0 | 6 (30%) | |
| | Other | 0 | 3 (15%) | |
| Contraception | COC | 5 (7%) | 9 (45%) | < 0.001 |
| | DMPA | 54 (77%) | 1 (5%) | |
| | NET-EN | 11 (16%) | 0 (0%) | |
| | Implant | 0 (0%) | 1 (5%) | |
| | Vaginal ring | 0 (0%) | 1 (5%) | |
| | Lng -IUD | 0 (0%) | 6 (30%) | |
| | Other | 0 (0%) | 2 (10%) | |
| Food insecurity | | 28 (40%) | 3 (15%) | 0.038 |
| Lifetime partners | | 4 (1-20) | 14 (4-90) | < 0.001 |
| Partners last month | | 1 (0-3) | 1 (0-2) | 0.4 |
| COC = Combined estrogen/progesterone oral contraceptives; DMPA = Depo medroxyprogesterone acetate; NET-EN = norethisterone enanthate; Implant = etonorgestrel contraceptive implant; Lng-IUD = levonorgestrel containing intrauterine device | | | | |

**Supplemental Table 3:** Sensitivity analysis using only weeks 3-5 to assess the primary outcome in the two arms with 7-day dosing.

| | Placebo | LC106-7 | LC106-3 | LC106-o | LC115 |
|---|---|---|---|---|---|
| South Africa | N = 14 | N = 14 | N = 14 | N = 14 | N = 14 |
| n (%) with outcome | 0 (0%) | 8 (57%) | 7 (50%) | 7 (50%) | 6 (43%) |
| 95% CI | | 29%-82% | 23%-77% | 23%-77% | 18%-71% |
| United States | N = 5 | N = 7 | N = 1 | N = 1 | N = 6 |
| n (%) with outcome | 1 (20%) | 4 (57%) | 1 (100%) | 1 (100%) | 4 (67%) |
| 95% CI | | 18% - 90% | 3% - 100% | 3% - 100% | 22% - 96% |
| RR (95% CI) | Ref | 10.86 (1.38 – inf) | 10.13 (1.37 – inf) | 10.13 (1.37 – inf) | 9.5 (1.3 – inf) |

**Supplemental Table 4:** Source of the individual strains used in the LBP, the number of chromosomal genes, presence of mobile genetic elements and their corresponding gene number.

| Strain | From | Number of genes |
|---|---|---|
| C0022A1 | White, Hispanic, Pregnant | 2491 |
| C0059E1 | Black, Non-Hispanic | 2773 |
| C0175A1 | Filipino, Pregnant | 2427 + plasmid w/20 genes |
| C00006A1 | White, non-Hispanic | 2333 + plasmid w/20 genes |
| C0028A1 | Hispanic | 2380 + plasmid w/171 genes |
| C0112A1 | White, non-Hispanic, Pregnant | 2477 |
| FF00004 | Black South African | 2503 + prophage w/75 genes |
| FF00018 | Black South African | 2518 + 2 plasmids |
| FF00051 | Black South African | 2554 |
| FF00064 | Black South African | 2466 |
| FF00072 | Black South African | 2577 + plasmid w/23 genes |
| UC101 | Black South African | 2588 |
| UC119 | Black South African | 2597 + plasmid w/21 genes |
| 185329 | Black South African | 2323 + plasmid w/46 genes |
| 122010 | Black South African | 2560 + plasmid w/12 genes |

**Supplemental Table 5:** Primers and probes for the strain-specific qPCR

| Group | Strain | Fluorophore | Forward (5'-3') | Reverse (5'-3') | Probe (5'-3') |
|---|---|---|---|---|---|
| 1 | C0175A1 | Cy5 | TGTTCCCGCCAATAATCCTAAA | CCCTACTAAACACAGATTTACTACCC | CCCTGCCCTAGTCAACGCTAGTTT |
| | C0022A1 | FAM | GGCAGCTTATGCTGATGAAATAC | GTAGTGTTTCCTACCGTAGCTTTA | TCGACAATCCTTCTGGAAATGGAAA |
| | C0059E1 | HEX | GCGTTAGAAGGTTTGGGTAAATC | GAGGGTAACTATTATCGTCATATCTTTG | AGGTGAACTAAGATTCAAAGGTGAGCT |
| 2 | UC101 | Cy5 | CCGCCACTGATATTGCTAAGA | TATTTGTCCAGCGGCCATAC | AGCTGAACAACCTGGGCAGAATGA |
| | FF00018 | FAM | GTGGGTAACGGACAACTCTTAT | TTACCACATCTCCCGGATCT | ATTGCATCCTACACAGAAGCCCGT |
| | FF00051 | HEX | AAGCGTGATAATGGTATCTGCTAT | TCCAAGTATGTGTTCCACCTTT | AGCCCAGATGCTCCTACAATTGAACA |
| 3 | 185329 | Cy5 | CCCAGCTTTCAGCTTTCATAATTT | GAATGTCATGGGACATGGTAGG | TCATTTGCTAAGCCATCTCTCAGCATCT |
| | C0028A1 | FAM | TCTTAGTGGAAATGGAGCAGAAA | CGAATACTTTGCATCAGCTTGATTA | ATCACAGCGACTGGAACCATCAGT |
| | C0112A1 | HEX | GGCAAACCGCTAACTTTGATTA | TTCTTGCTGTTAGGATTAACGATTAC | ACTTTGTTAAGCTGGCTAAGTCCGA |
| 4 | FF00004 | Cy5 | CCCAATGGTACGAACCTCAA | GAGGCATATGAGCAATTCAACAA | CCTACGGCTTCCGCTAATTGCTCT |
| | C0006A1 | FAM | AATGTTGGAAATGACCCTGTTTC | CAACACCAAATGCGGCATAC | GGATCACTCAACTGATCCCGCCAA |
| | 122010 | HEX | CTTTGTAAGGAATATGCCGTTCTG | GTACTGGCACAGATGTCGTAG | TTTAGTTGACCGTGGTGTCATGGG |
| 5 | UC119 | Cy5 | ATGACTAGTGAACCAGTAACTT | CTTTAGCTTGGTTATGGTTTGT | ACGGCTTAATTACCGCTGATGGCT |
| | FF00064 | FAM | TCTCGTCAATCAGTTTCTAAATGGG | AGATGGGTAGAGGGCGATTT | AGGTGGTTCTTTACCAGATATTGACCGT |
| | FF00072 | HEX | GGATCACAATGACGTACCTTACA | TCGTCACCATCGTAAGTATCATTT | AGCAAATGATCCCAACCGTAAGCCT |
